# Estimating the risk on outbreak spreading of 2019-nCoV in China using transportation data

**DOI:** 10.1101/2020.02.01.20019984

**Authors:** Hsiang-Yu Yuan, M. Pear Hossain, Mesfin Tsegaye, Xiaolin Zhu, Pengfei Jia, Alvin Junus, Tzai-Hung Wen, Dirk Pfeiffer

**Affiliations:** Department of Biomedical Sciences, Jockey Club College of Veterinary Medicine and Life Sciences, City University of Hong Kong, Hong Kong; Department of Statistics, Bangabandhu Sheikh Mujibur Rahman Science and Technology University, Bangladesh; Department of Land Surveying and Geo-Informatics, The Hong Kong Polytechnic University, Hong Kong; Academic Information Center, China Academy of Urban Planning and Design, Beijing, China; Department of Geography, National Taiwan University, Taiwan; Centre for Applied One Health Research and Policy Advice, City University of Hong Kong, Hong Kong

## Abstract

A novel corona virus (2019-nCoV) was identified in Wuhan, China and has been causing an unprecedented outbreak in China. The spread of this novel virus can eventually become an international emergency. During the early outbreak phase in Wuhan, one of the most important public health tasks is to prevent the spread of the virus to other cities. Therefore, full-scale border control measures to prevent the spread of virus have been discussed in many nearby countries. At the same time, lockdown in Wuhan cityu (border control from leaving out) has been imposed. The challenge is that many people have traveled from Wuhan to other cities before the border control. Thus, it is difficult to forecast the number of imported cases at different cities and estimate their risk on outbreak emergence.

Here, we have developed a mathematical framework incorporating city-to-city connections to calculate the number of imported cases of the novel virus from an outbreak source, and the cumulative number of secondary cases generated by the imported cases. We used this number to estimate the arrival time of outbreak emergence using air travel frequency data from Wuhan to other cities, collected from the International Air Transport Association database. In addition, a meta-population compartmental model was built based on a classical SIR approach to simulate outbreaks at different cities.

We consider the scenarios under three basic reproductive number (*R*_0_) settings using the best knowledge of the current findings, from high (2.92), mild (1.68), to a much lower numbers (1.4). The mean arrival time of outbreak spreading has been determined. Under the high *R*_0_, the critical time is 17.9 days after December 31, 2019 for outbreak spreading. Under the low *R*_0_, the critical time is between day 26.2 to day 35 after December 31, 2019. To make an extra 30 days gain, under the low *R*_0_ (1.4), the control measures have to reduce 87% of the connections between the source and target cities. Under the higher *R*_0_ (2.92), the effect on reducing the chance of outbreak emergence is generally low until the border control measure was enhanced to reduce more than 95% of the connections.

## Introduction

On 31 December 2019, WHO was alerted to several cases of pneumonia infections in Wuhan City, Hubei Province of China [1]. The cause of the pneumonia was later identified as a novel Coronavirus (2019-nCoV) genetically very closely related to the Middle Eastern Respiratory Syndrome virus (MERS-CoV) and the Severe Acute Respiratory Syndrome virus (SARSCoV) [2]. This marks the third time in 20 years that a member of the family of coronaviruses (CoVs) has caused an epidemic employing its zoonotic potential, for example, from bats [3]. The novel virus was able to establish between person-to-person transmission [4].

The outbreak caused by the novel coronavirus (2019-nCoV) is currently occurring in Wuhan and affecting many neighboring cities and countries [5]. During the early phase of the outbreak, it poses a severe threat to many other regions due to frequent transportation services linking Wuhan to the other cities. About 2 weeks later after 31 December, using the number of cases detected outside China, it has been inferred that more than a thousand individuals (with an estimated mean 1723) had had an onset of symptoms by January 12, 2020 [6]. This number, which was estimated based on certain assumptions, was about 40 times higher than the reported number by January 12, 2020. Given that most of the members of the coronavirus family cause mild flu-like symptoms during infection and most people have not yet received nation-wide alert about the novel virus during that period, it could be possible that the confirmed case number among actual infected cases is very low, probably similar to influenza reporting rate, which is only about 1% [7].

During this early outbreak phase in Wuhan, one of the most important preventive tasks is to prevent the outbreak potential to spread to other cities. If outbreaks also occur in other cities, the prevention of nCoV becomes even more difficult in China or at the global level. From disease control perspective, the major questions here are whether we can i) estimate the waiting time for nCoV to spread to other cities in China; and ii) evaluate the impacts of disease control by cessation of population movement (e.g., lockdown measures) on the outbreak spreading. The importance of prediction of infectious diseases based on transportation network information has already been highlighted previously [8, 9]. Studies have been done to estimate the probability of emergence and the arrival time of the emergence [10]. In addition, the required condition for a virus to spread to other cities is through secondary infections from the imported cases. In our study, imported case was defined as an infected individual migrating from other cities to a particular city. In order to evaluate the infectious disease control measures during the spreading of the virus, it is important to know how many secondary cases has been caused by the imported cases. Thus, there is a need to estimate the probability of emergence and the arrival time of the emergence through estimating the number of secondary infections caused from imported cases using transportation data.

In this study, we derived a simple mathematical formula to estimate the outbreak potential at neighboring cities and built a meta-population model based on a classical SIR approach to understand the outbreak spreading dynamics at different cities. Gain of time before outbreak emergence is predicted under different infectious disease control rates.

## Methods

### Calculating Imported & their secondary infected cases

Assuming that the newly emergence of 2019-nCoV causes an outbreak at location *i*, during the emergence, the number of infectious cases is changing following this formula:

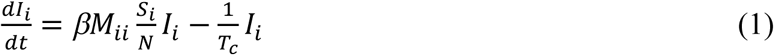

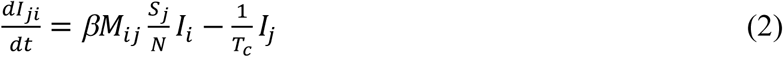

where *β* is the baseline transmission rate that can be estimated from 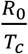, *R* is the basic reproductive number, *T*_*c*_ is the generation time, *M*_*ij*_ is the contact mixing from the location *i* to *j*, and *I*_*ji*_ is the secondary infected at the location *j* transmitted by imported cases from *i*. During the early outbreak phase, the susceptible population (*S*) is nearly constant, therefore we can assume 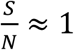, where *N* is total population size. Set 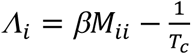, then 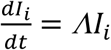 and we will have the infected number at *i* as a function of time *t*, such that *I*_*i*_(*t*) = *I*_0_*e*^*Λt*^. Because before the outbreak occurs at a different location *j*, the infection can only happen when transmission events occur between the imported cases and the individuals at *j*, it is essential to estimate the expected number of cumulated secondary cases in the population *j* transmitted from the imported cases originally at *i*. To solve the number of cumulated cases until time *t*, we have the number of cumulated secondary infected cases generated by the imported cases 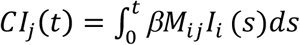. We can derive the following formula:

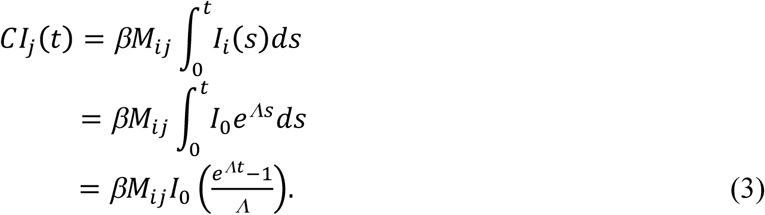

The total number of the imported cases during a period of time is derived as: 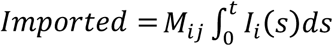. Note that we can incorporate the effect of latent period. For example, if we consider infected people during latent state or within incubation period can pass the border screening, during each time point *s*, only newly infected indiviauls within inbucation period τ can have chance to move to a different city, thus 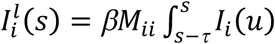. The total number of imported cases at a specific time point can also be derived as 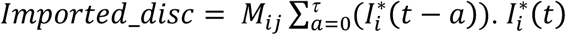 is the daily new infected, which can be calculated as *I*_*i*_ (*t*) − *I*_*i*_ (*t* − 1). We adopt a discrete form here because it is easier to explain. *I*_*i*_(*t* − *a*) represents the number of new cases that were infected *a* days ago but will still not be detected at time *t*. If we assume *I*_*i*_(*t* − *a*) is constant for all possible *a* and τ is the time to disease detection for each infected individual, the result is exactly same as the total number of imported cases estimated in a recent study [6]. Therefore, this framework provides a more generalized expression to estimate the imported cases and the secondary infected cases generated by the imported cases when the the incidence is still exponentially increasing in the source region.

### Incorporation border control measures

To incorporate the border control measure into our model, we introduce a control factor *c* such that

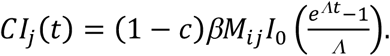

where *c* is the border control measure ranging from 0 to 1, representing no control to full-scaled control.

### Determining outbreak potentials

We consider the outbreak potential, defined as the probability of outbreak emergence given the number of cumulated cases. At the initial stage, if there is only one single infected individual, the chance of this virus to cause an outbreak is 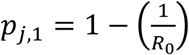. We set *R*_0_ for location *j* to be a low number 1.1 for the secondary infected cases generated by the imported cases at location at *j* because the nation-wide alert has already been received at different cities after December 31, 2019. When the number of cumulative cases reaches a higher number, the probability of outbreak emergence is

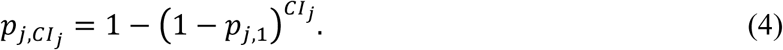

We obtain a critical threshold number of cases, 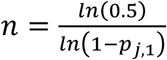, representing the threshold of having higher outbreak probability. We set the critical time *t*_*d*_ such that the probability of outbreak emergence 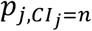 is larger than 50% can be solved.

Airline frequency data is collected from the International Air Transport Association (IATA) database. Although we do not have data for rail and other forms of transport, we assumed that the total number of travelers is 3.37 times higher than that of air transport except for certain cities on Hainan island. The total travel volume between different cities was then used for the contact mixing *M*_*ij*_ between locations *i* and *j*. For example, we divided daily passenger numbers by total population size in Wuhan to represent the contact rate between Wuhan city and any connected city *j*.

## Results

### Outbreak spreading during early phase in a higher R_0_ scenario

We first calculated the cumulative number of secondary infected cases by import cases from Wuhan under two parameter settings, each corresponding to a higher or lower *R*_0_. For higher *R*_0_ scenario, a generation time of 8.4 days [11] and *R*_0_ of 2.92 for the outbreak in Wuhan city were used [12]. We predicted the border control impacts on nation-wide disease spreading in two different scenarios, without any other control and with some control measures. During the early stage of the outbreak without any control measures (no mask wearing, isolation, or contact tracing, etc.), the cumulative number of secondary infections generated by imported cases through transportation was estimated (Figure 1A). We obtain the critical threshold number of cases *n* = 7.27 in a city, representing the outbreak will occur with 50% chance if the cumulative number of secondary infections is over that critical value. The arrival time of secondary infections to reach above this value for the top 10 cities most connected to Wuhan are between 14 – 18 days (Figure 1A). The outbreak potential in the nearby cities can be assessed. Using the average of the airline frequencies among the 10 top connected cities, we calculated the average arrival time of outbreak emergence 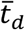, which is 17.9 days after 31 December 2019. At the time 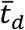, all the top 10 connected cities have a probability of outbreak emergence larger than 50% (Figure 1B).

**Figure 1.**
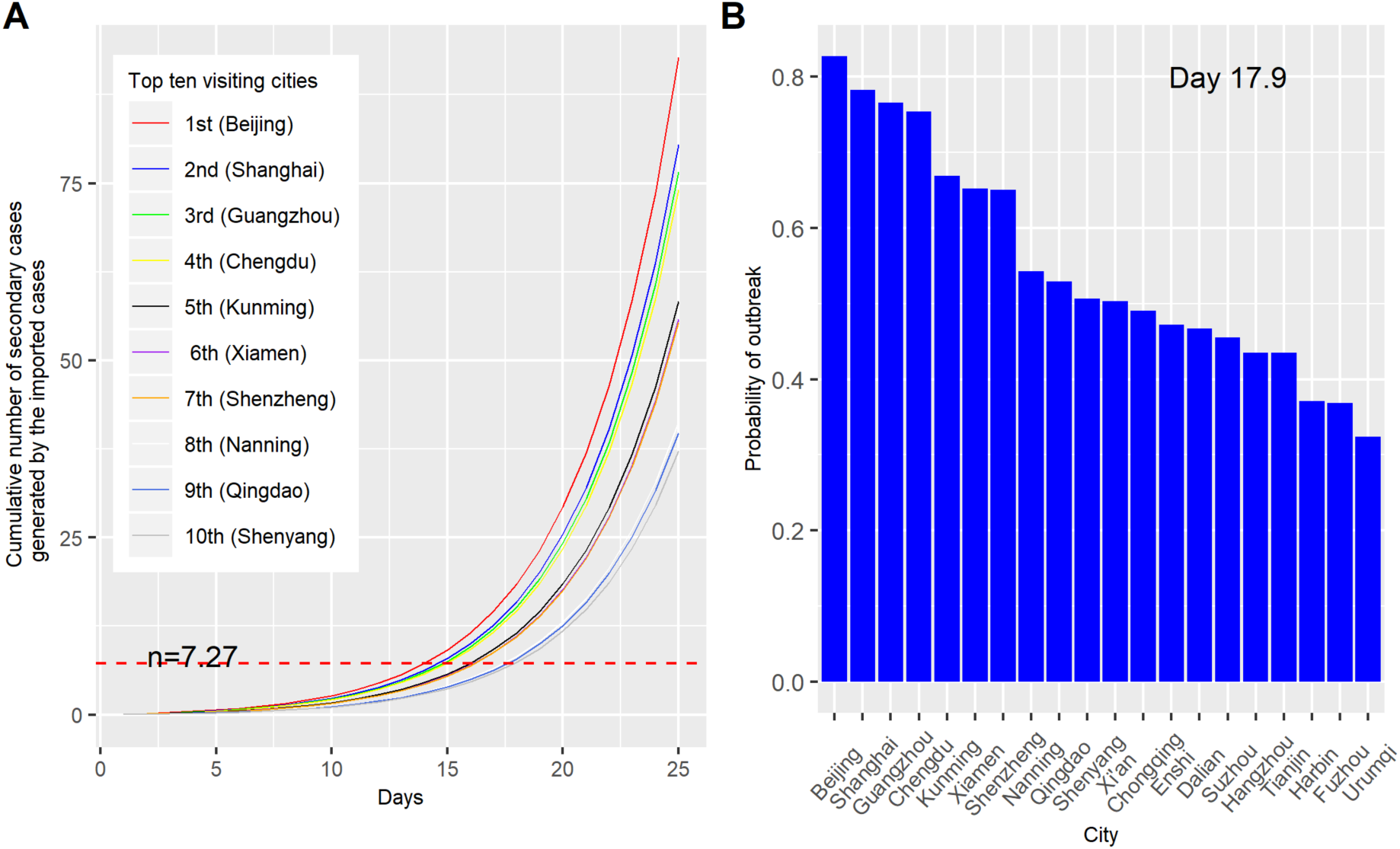
Outbreak potential estimated from the secondary cases contacted by imported cases under a higher *R*” scenario. (A) Number of cumulative secondary cases generated by imported cases. The secondary infecteds are listed among the top 10 visiting cities from Wuhan. *n* = 7.27 is the critical threshold number. (B) Probability of outbreak emergence in different cities at mean critical time (17.9 days).

For each city, the critical time at which the cumulative number of secondary cases generated by the imported cases larger than the critical threshold were determined (Table 1). Beijing, Shanghai, Guangzhou and and Chengdu had the shortest required time periods. As stated before, the mean critical time for the top 10 visiting cities was 17.9 days. As shown in Figure 1, only eleven cities had higher risk of outbreak emergence (probability larger than 0.5).

**Table 1.** Critical time of outbreak arrival at top 20 connected cities in China under a higher *R*_0_. *CI*_*j*_ is the cumulated number of secondary infected cases generated by the imported cases.

| City | Critical time (days) | $CI_j$ at mean critical time (day 17.9) | $CI_j$ at day 35 | Probability of outbreak at mean critical time (day 17.9) |
| --- | --- | --- | --- | --- |
| Beijing | 15 | 18.4 | 918.1 | 0.827 |
| Shanghai | 15 | 16.0 | 796.7 | 0.782 |
| Guangzhou | 15 | 15.2 | 758.3 | 0.766 |
| Chengdu | 15 | 14.7 | 732.8 | 0.754 |
| Kunming | 17 | 11.6 | 577.3 | 0.669 |
| Xiamen | 17 | 11.1 | 552.3 | 0.652 |
| Shenzhen | 17 | 11.0 | 548.7 | 0.650 |
| Nanning | 18 | 8.2 | 408.7 | 0.543 |
| Qingdao | 18 | 7.9 | 394.0 | 0.529 |
| Shenyang | 18 | 7.4 | 368.9 | 0.506 |
| Xian | 18 | 7.3 | 365.9 | 0.503 |
| Chongqing | 19 | 7.1 | 352.8 | 0.491 |
| Enshi | 19 | 6.7 | 334.1 | 0.472 |
| Dalian | 19 | 6.6 | 329.0 | 0.467 |
| Suzhou | 19 | 6.4 | 317.7 | 0.456 |
| Hangzhou | 19 | 6.0 | 298.7 | 0.435 |
| Tianjin | 19 | 6.0 | 298.1 | 0.435 |
| Harbin | 20 | 4.9 | 242.2 | 0.371 |
| Fuzhou | 20 | 4.8 | 240.6 | 0.369 |
| Urumqi | 21 | 4.1 | 204.6 | 0.324 |

### Outbreak spreading during early phase in a lower R_0_ scenario

In reality, the *R*_0_ or real-time effective *R* will decrease after many control measures are conducted [13]. Therefore, we made the same calculation under a mild transmission condition. We assumed that after 31 December 2019, the generation time was as short as 7 days and *R*_0_ was as low as 1.4, which was the lower bound of World Health Organization’s estimate [14]. The lower *R*_0_ can be caused by different estimation procedure or can be caused by isolation and mask wearing measures. Under this scenario, following the suggestion from a recent study [15], we consider the reporting rate to be near 5% and set initial infected number as 1000 on 31 December. Using model simulation, the infected number reached the recent estimated number 1723 on 9 January 2020 (Figure 2). Considering the incubation period to be 3 days, the number of infected persons with symptoms onset will be revealed on 12 January, 2020, same as the value estimated from the study [6]. The infected number was slowly increasing linearly within the first 30 days. In the following study, we use this mild *R*_0_ parameter setting to evalulate the impact of border control on outbreak spreading.

**Figure 2.**
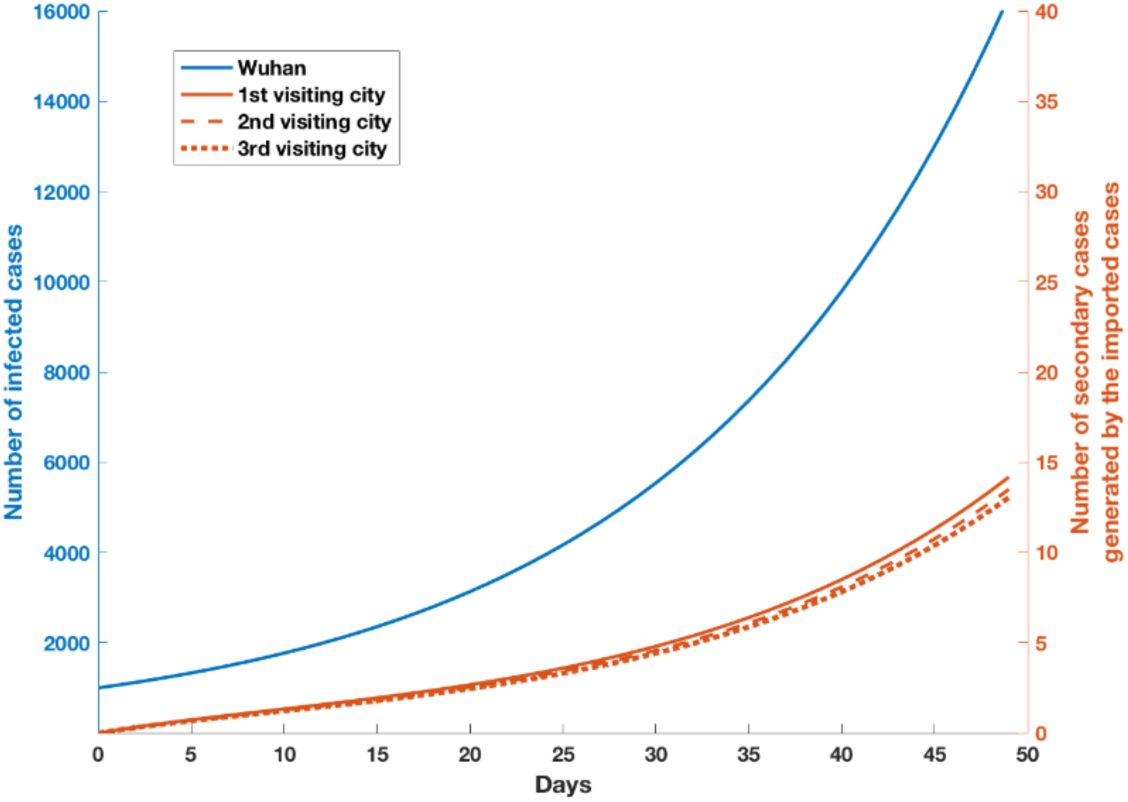
Number of infected cases and number of secondary cases generated by the imported cases. The blue line represents the number of infected cases of nCoV after 31 December 2019 and the orange lines represent the number of secondary cases generated by the imported cases through contact between cross-province transportation.

We further calculated the cumulative number of secondary cases *CI*_*j*_ at the *j*th city generated by the imported cases. The changes of *CI*_*j*_ by time at each city are plotted from day 1 to 50 (Figure 3A). After 50 days since the outbreak, the cumulative cases can reach rapidly above 40 cases for many cities. After 30 days, most of the cities have secondary cases above a critical threshold of 7.27, calculated from eq(4) using *R*_0_ = 1.1. The current estimate of *R*_0_ is 2.92 for the initial outbreak period in Wuhan when citizens had not received any alert. However, the current *R*_0_ would be lower because people have since been aware of the disease. This setting will apply when about 62.3% of infections events can be prevented, due to different control policies, comparing to the Wuhan outbreak before the emergence of outbreak on other cities.

**Figure 3.**
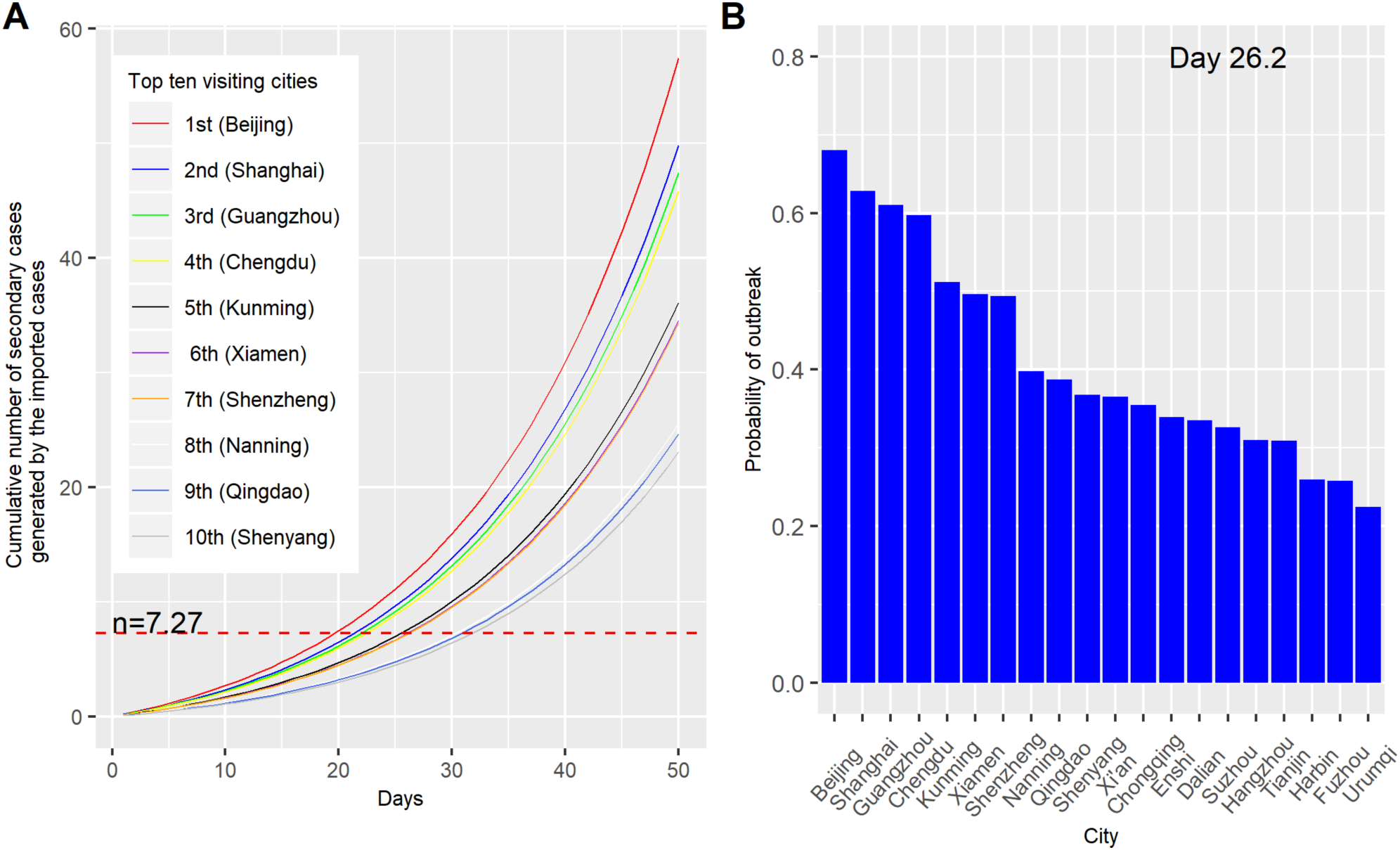
Outbreak potential estimated from the secondary cases contacted by imported cases under a lower *R*” scenario. (A) Number of cumulative secondary cases generated by imported cases. The secondary infecteds are listed among the top 10 visiting cities from Wuhan. *n* = 7.27 is the critical threshold number. (B) Probability of outbreak emergence in different cities at mean critical time (26.2 days).

The outbreak potential of nearby cities can be assessed. For each city, the critical time at which the cumulative number of secondary cases generated by the imported cases larger than the critical threshold was determined (Table 2). Beijing, Shanghai, Guangzhou had the shortest required time periods. The mean critical time from the top 10 visiting cities was 26.2 days. Only five cities had higher risk of outbreak emergence (probability larger than 0.5). We further calculated the probability of outbreak emergence at mean critical time and at day 35. 17 cities had higher risk of outbreak emergence (Table 2). We further compared the predicted cumulated infected case number at day 26 using model simulation with the actual observed reports in the top 10 visitng cities. Among top 5 high risk cities, four of them had higher case numbers than other cities. The higher number of predicted numbers confirmed that the actual case number can be 20 times higher than the reported numbers (*see suplemnetary* Table S1).

**Table 2.**
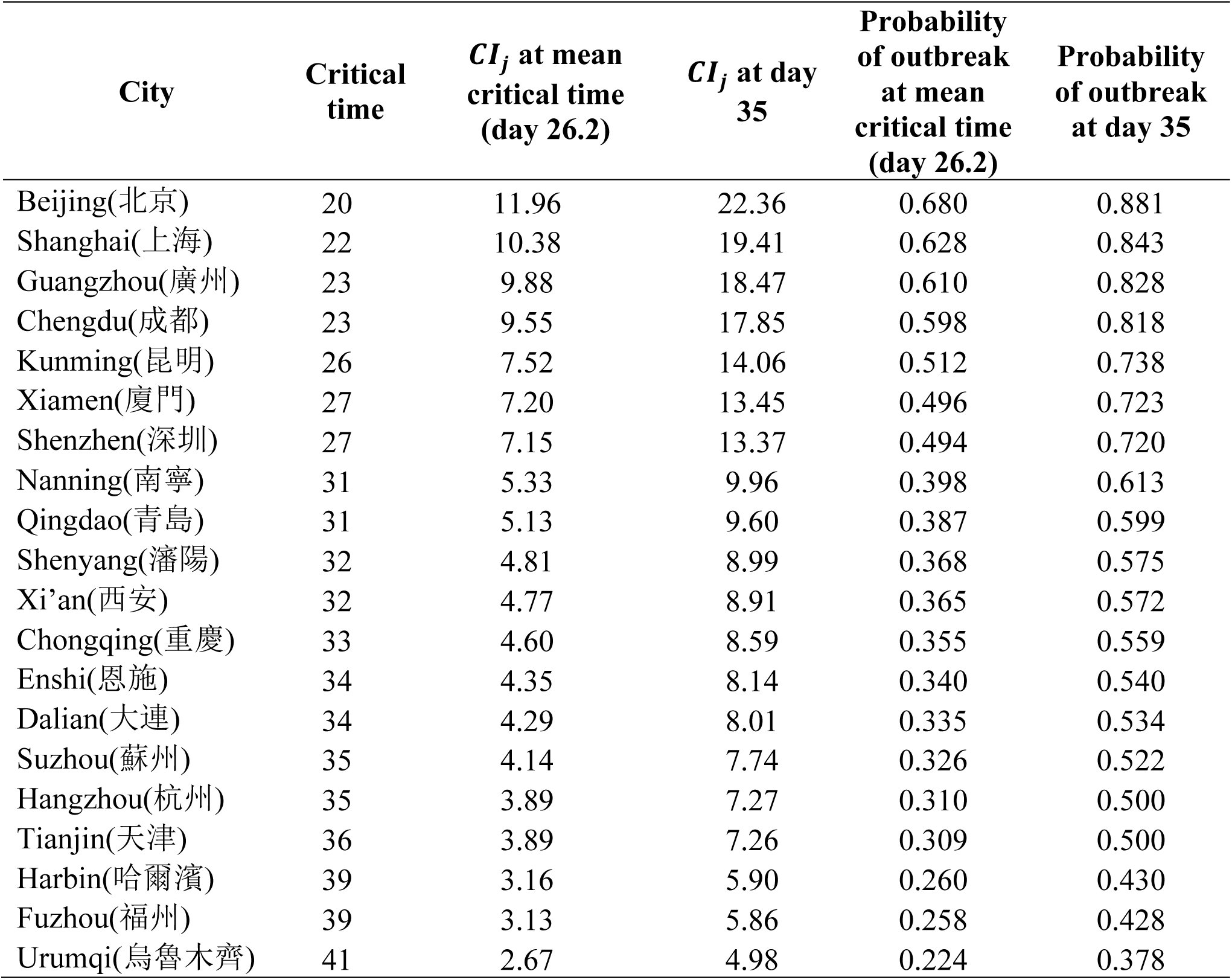
Critical time of outbreak arrival at top 20 frequently-visited cities in China under a lower *R*_0_. *CI*_*j*_ is the cumulated number of secondary infected cases generated by the imported cases.

| City | Critical time | $CI_j$ at mean critical time (day 26.2) | $CI_j$ at day 35 | Probability of outbreak at mean critical time (day 26.2) | Probability of outbreak at day 35 |
| --- | --- | --- | --- | --- | --- |
| Beijing(北京) | 20 | 11.96 | 22.36 | 0.680 | 0.881 |
| Shanghai(上海) | 22 | 10.38 | 19.41 | 0.628 | 0.843 |
| Guangzhou(廣州) | 23 | 9.88 | 18.47 | 0.610 | 0.828 |
| Chengdu(成都) | 23 | 9.55 | 17.85 | 0.598 | 0.818 |
| Kunming(昆明) | 26 | 7.52 | 14.06 | 0.512 | 0.738 |
| Xiamen(廈門) | 27 | 7.20 | 13.45 | 0.496 | 0.723 |
| Shenzhen(深圳) | 27 | 7.15 | 13.37 | 0.494 | 0.720 |
| Nanning(南寧) | 31 | 5.33 | 9.96 | 0.398 | 0.613 |
| Qingdao(青島) | 31 | 5.13 | 9.60 | 0.387 | 0.599 |
| Shenyang(瀋陽) | 32 | 4.81 | 8.99 | 0.368 | 0.575 |
| Xi'an(西安) | 32 | 4.77 | 8.91 | 0.365 | 0.572 |
| Chongqing(重慶) | 33 | 4.60 | 8.59 | 0.355 | 0.559 |
| Enshi(恩施) | 34 | 4.35 | 8.14 | 0.340 | 0.540 |
| Dalian(大連) | 34 | 4.29 | 8.01 | 0.335 | 0.534 |
| Suzhou(蘇州) | 35 | 4.14 | 7.74 | 0.326 | 0.522 |
| Hangzhou(杭州) | 35 | 3.89 | 7.27 | 0.310 | 0.500 |
| Tianjin(天津) | 36 | 3.89 | 7.26 | 0.309 | 0.500 |
| Harbin(哈爾濱) | 39 | 3.16 | 5.90 | 0.260 | 0.430 |
| Fuzhou(福州) | 39 | 3.13 | 5.86 | 0.258 | 0.428 |
| Urumqi(烏魯木齊) | 41 | 2.67 | 4.98 | 0.224 | 0.378 |

### Probability of outbreak emergence and spreading

Under this parameter setting, the critical time *t*_*d*_ (arrival time for emergence) when the probability of outbreak emergence at different cities larger than 0.5 were calculated. The probability of outbreak emergence at each of the top 20 frequently-visited cities at the average 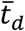 (Figure 4A) and day 35 (Figure 4B) were plotted.

**Figure 4.**
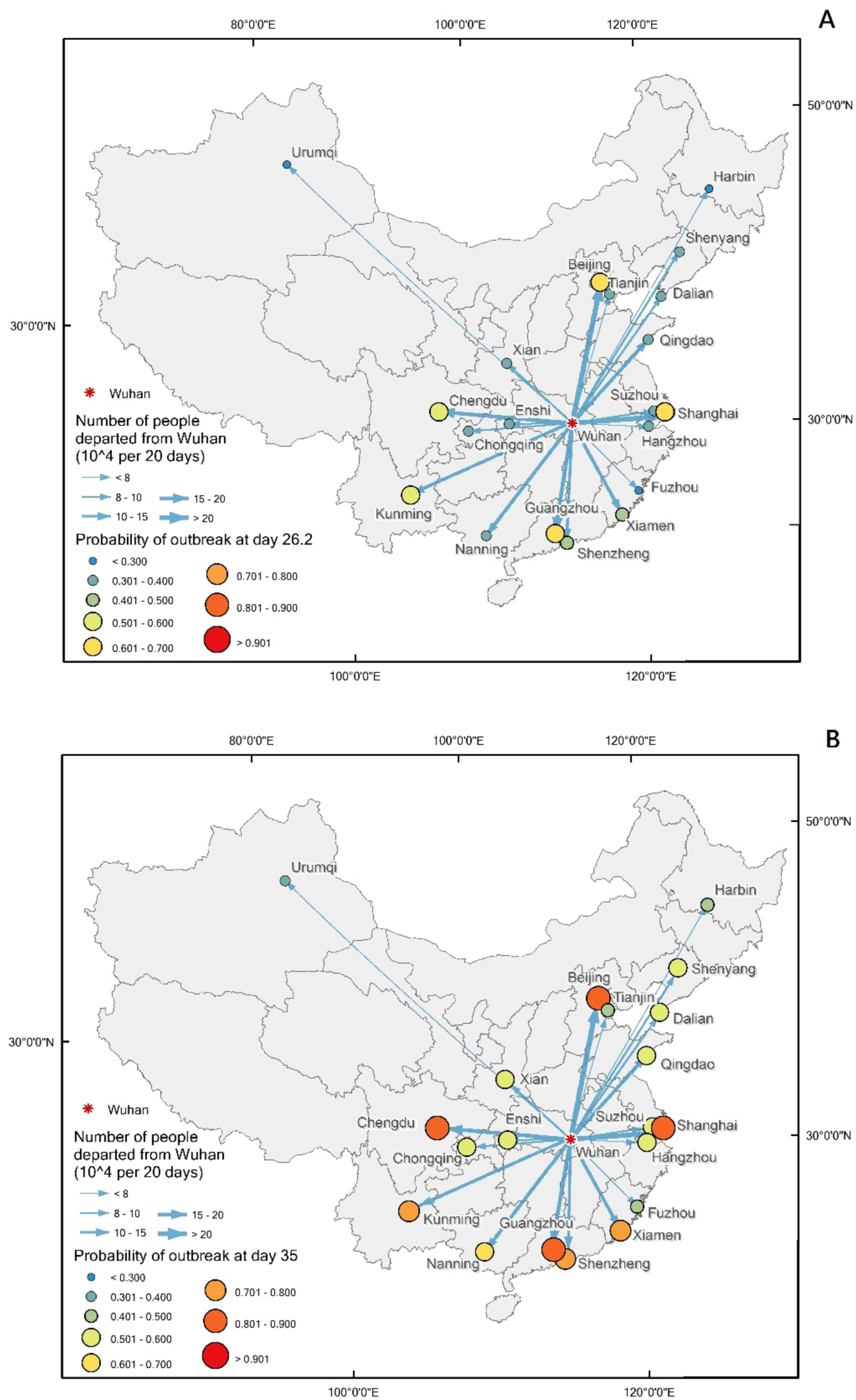
Outbreak potential at different cities in mainland China. Outbreak potential at different cities at day 26.2 (A) and day 35 (B).

### Evaluation of the impacts of border control

Under the no-border control scenario, assuming all other cities have the same population as Wuhan, model simulation results showed that nearly 2 months were required for generating the same level of epidemics in other frequent visiting cities (Figure 5). Ideally, complete cessation of population movement between cities or isolation of every susceptible case from the source city can reduce transmission events.

**Figure 5.**
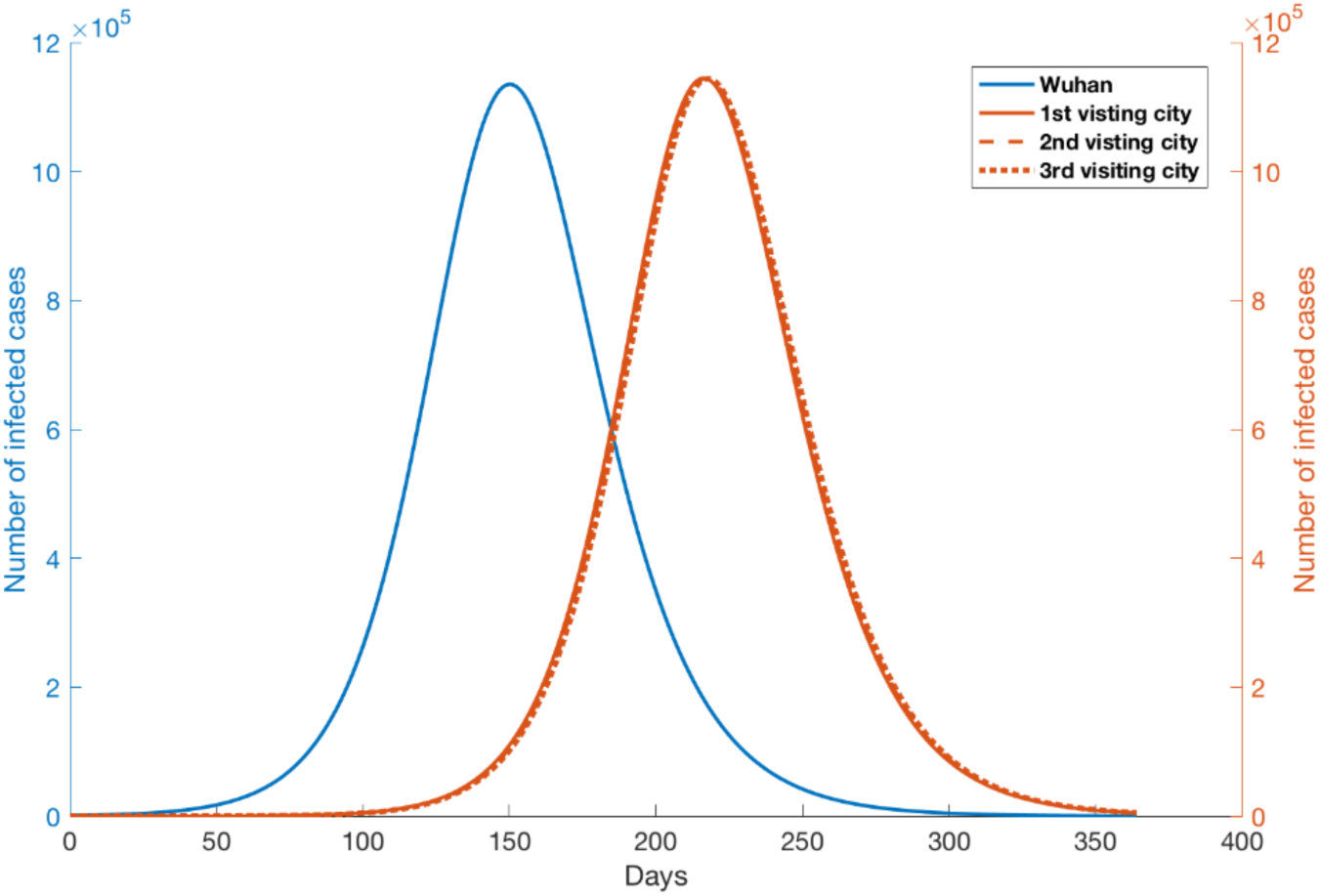
Epidemic outbreaks under no disease control scenario. Blue line represents the number of infected cases from Wuhan city. Orange lines represent the number of infected cases from top 3 visiting cities from Wuhan.

We evaluated the impact of border control measures by introducting a control rate *c* to reduce the transporation rate between two cities, such that 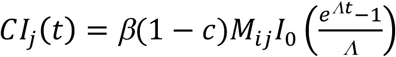. We can thus recalculate the cumulative number of secondary infections under different levels of border control. To achieve the gain time of control for transportation of a particular city, we calculate critical time given the control such that the cumulative number of secondary cases generated by the imported cases is greater than critical threshold number *n*. Finally, subtraction of the critical time without control from the critical time with control returns gain of the control system. To make an extra 30 days gain, under the low *R*_0_ (1.4), the control measures have to reduce 87% of the secondary infections generated by the imported cases (Figure 6A). Under the higher *R*_0_ (2.92), the effect on reducing the chance of outbreak emergence is low until the border control can reduce more than 95% of the secondary infections (Figure 6B). Under the mild *R*_0_ condition, we set *R*_0_ = 1.6*c* with generation time 8.4 days, and found that the border control effect is similar but a little bit weaker than that in the lower *R*_0_ scenario. This allows us to calculate how much extra time a connected region through mobility can gain and gives an indicator what scale of border control is necessary.

**Figure 6.**
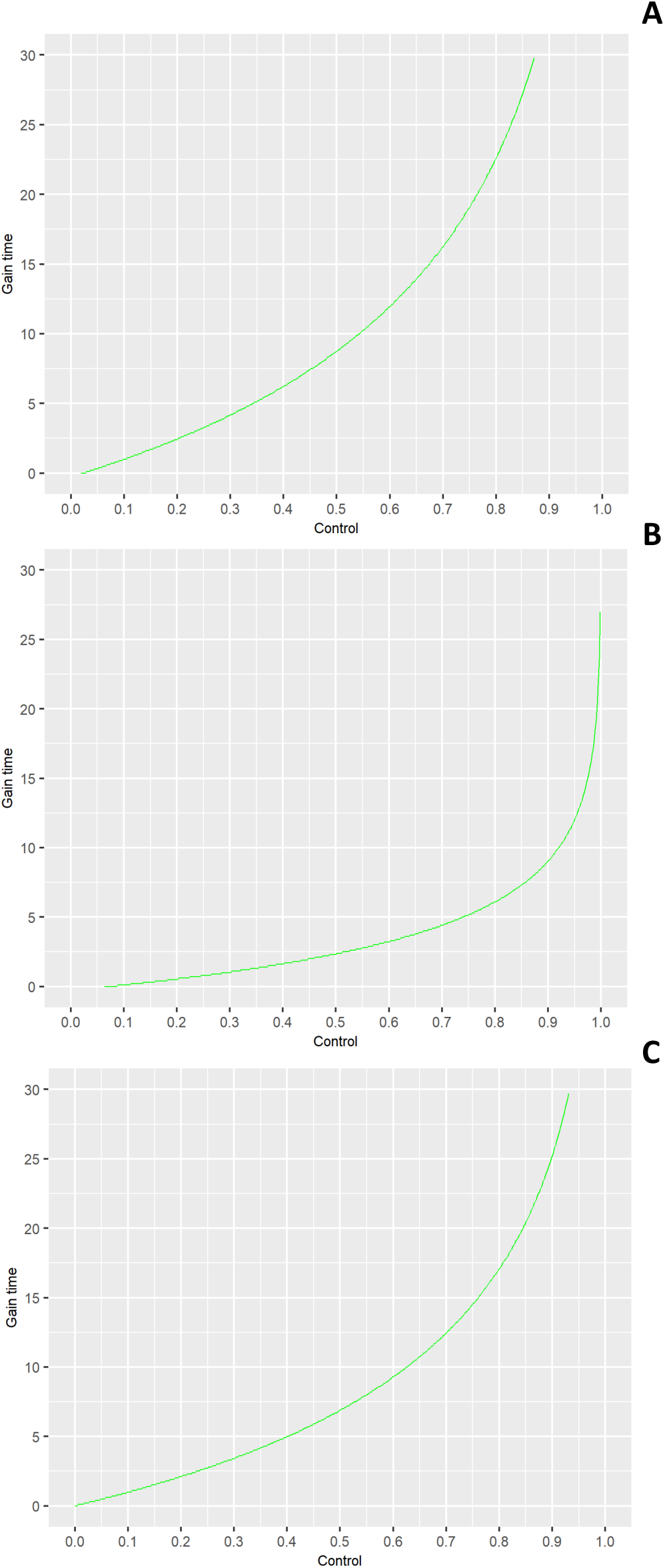
Gain time of outbreak emergence by the rates of successful infectious disease border control. (A) The impact of border control under the lower *R*_0_ (1.4); (B) under the higher *R*_0_ (2.92); (C) under mild *R*_0_ (1.68).

## Discussion

The 2019-nCoV is currently spreading from Wuhan to many nearby cities and countries. It is hypothesized that the rate of transmission between different cities or countries is proportional to the number of people moving from different locations to Wuhan. Until recently, very few studies have been done to predict the spreading of the outbreaks of newly emerging diseases using air and other forms of transport information. Our study provides an analytical solution to calculate the cumulative number of secondary cases generated by the imported cases with an estimated *R*_0_, with which we are able to predict the arrival time of the outbreak.

Two most important parameters, generation time and basic reproduction number are still unknown. The generation time we used was adapted from a previous study. Since *R*_0_ is not known, and can be different using different approaches [16], here we use *R*_0_ = 1.4 for Wuhan and we set *R*_0_ for other locations *j* a low number 1.1. This number was used because the nation-wide alert has already been received at different cities after 31 December, 2019. The control measures and self awareness make the initial infection to occur less easily. We have learnt from the previous SARS outbreak that it is crucial to implement rapid infection control measures to limit the impact of epidemics, both in terms of preventing more casualties and shortening of the epidemic period. The other important lesson is that a timely public alert against a new spreading epidemic is also most essential [17].

Previous study shows that control measures at international cross-borders and screening at borders have influential effect in mitigating the spread of infectious diseases [18]. They strongly recommend to strengthen cross-boarder screening system to prevent infectious disease outbreak due to imported cases as China has increased number of imported cases over the years. Besides, [19] studied the effectiveness of border screening for influenza detection among airline passengers in New Zealand. Here the model we constructed is easy to understand and simple to use when transportation data is available. The framework can be extended to multiple infected sources to one single target city. Hence, the model proposed in the current study could help to prevent infectious diseases caused by imported cases from different parts of China moving to neighbouring territories and countries including Hong Kong, Taiwan, Vietnam and so on.

Ideally, complete cessation of population movement between cities will be a way to block outbreak spreading. However, the effect is difficult to measure given that it is not easy to make a comparative study. But a lesson we learnt from the previous SARS outbreak is that it is crucial to estimate the transmission potential of a new emerging disease as soon as possible and to establish whether additional, more stringent control measures are required [17]. Recent studies have shown the importance of modeling in infectious disease control [20]. We have evaluated the impact of these control measures under different levels of infectious disease control scenarios. To make an extra 30 days gain, the control measures have to reduce 87% of secondary infections generated by the imported cases. This gives an indicator whether certain control or prevention measures, such as wearing masks, quarantine, etc., are necessary.

## Data Availability

Data are available on request.

## Acknowledgements

We thank Miss Xiaoyue Tan for her assistance in making maps. We thank Prof. Mengsu (Michael) Yang and all the anonymous readers to who have provided invaluable comments. We thank for funding supports from City University of Hong Kong (#7200573 and #9610416).

## Author contributions

HY and XZ designed the study. PJ participated in the data collection. HY, MPH, XZ, PJ and AJ analysed and interpreted the data. HY, MT and MPH wrote the paper. Everyone reviewed, revised and edited the manuscript.

